# Self-monitoring of blood glucose levels among pregnant individuals with gestational diabetes: a systematic review and meta-analysis

**DOI:** 10.1101/2022.08.11.22278238

**Authors:** Ping Teresa Yeh, Caitlin Elizabeth Kennedy, Dong Keun Rhee, Chloe Zera, Özge Tunçalp, Briana Lucido, Rodolfo Gomez Ponce de Leon, Manjulaa Narasimhan

## Abstract

1

**Introduction:** The World Health Organization (WHO) recommends treatment and management of gestational diabetes (GD) through lifestyle changes, including diet and exercise, and self-monitoring blood glucose (SMBG) to inform timely treatment decisions. To expand the evidence base of WHO’s guideline on self-care interventions, we conducted a systematic review of SMBG among pregnant individuals with GD.

**Setting:** We searched for publications through November 2020 comparing SMBG with clinic-based monitoring during antenatal care (ANC) globally.

**Primary and secondary outcome measures:** We extracted data using standardized forms and summarized maternal and newborn findings using random effects meta-analysis in GRADE evidence tables. We also reviewed studies on values, preferences, and costs of SMBG.

**Results:** We identified 6 studies examining SMBG compared to routine ANC care, 5 studies on values and preferences, and 1 study on costs. Nearly all were conducted in Europe and North America. Moderate-certainty evidence from 3 randomized controlled trials (RCTs) showed that SMBG as part of a package of interventions for GD treatment was associated with lower rates of preeclampsia, lower mean birthweight, fewer infants born large for gestational age, fewer infants with macrosomia, and lower rates of shoulder dystocia. There was no difference between groups in self-efficacy, preterm birth, C-section, mental health, stillbirth, or respiratory distress. No studies measured placenta previa, long-term complications, device-related issues, or social harms. Most end-users supported SMBG, motivated by health benefits, convenience, ease of use, and increased confidence. Health workers acknowledged SMBG’s convenience but were wary of technical problems. One study found SMBG by pregnant individuals with insulin-dependent diabetes was associated with decreased costs for hospital admission and length of stay.

**Conclusion:** SMBG during pregnancy is feasible and acceptable, and when combined in a package of GD interventions, is generally associated with improved maternal and neonatal health outcomes. However, research from resource-limited settings is needed.

**Systematic review registration number:** PROSPERO CRD42021233862

## 2.1 Introduction

Gestational diabetes (GD) is defined as glucose intolerance resulting in clinical hyperglycemia with onset or first recognition during pregnancy.[1, 2] Hyperglycemia during pregnancy is associated with adverse short-term and long-term maternal and newborn health outcomes. Self-management of GD through lifestyle modification, including diet and exercise, is considered first-line treatment by health workers and several professional associations.[3-5] One component of GD self-management is self-monitoring of blood glucose (SMBG) levels, which is used clinically to monitor the effectiveness of lifestyle changes, guide intensification of treatment, and inform ANC.

This systematic review sought to examine the evidence for SMBG compared with monitoring of blood glucose levels by health workers within the ANC (clinic) setting. We conducted this systematic review in the context of expanding the evidence base of the World Health Organization (WHO) guideline on self-care interventions for health,[6] which includes several recommendations on self-care interventions during pregnancy, childbirth and post-natal care.[7, 8] WHO’s 2020 “Package of Essential Noncommunicable Disease Interventions for Primary Health Care” recommends ‘non-pharmacological’ treatment for management of type 2 diabetes.[9] This could be considered self-care, though it does not specify how/by whom diabetes should be diagnosed or monitored. Furthermore, self-monitoring may be a feasible approach when health services are disrupted such as in emergency or humanitarian settings. In the context of maintaining essential health services during the COVID-19 pandemic, WHO recommends creation of self-management plans for diabetes, if appropriate, supported by health workers.[10]

## 2.2 Methods

This review addressed the following overarching question: Should SMBG among pregnant women and other pregnant people^1^ with GD be made available in addition to clinic check-ups? Per the WHO guideline development process which incorporates multiple aspects of evidence,[11] we reviewed the extant literature in three areas relevant to this question: effectiveness of the intervention (what is the impact on the outcomes of interest when comparing SMBG to glucose monitoring at clinic check-ups for pregnant individuals with GD?), values and preferences of end users and health workers (what do patients and health workers think of SMBG?), and cost information (what are the costs (to the patient and to the health system) of SMBG?).

We followed Preferred Reporting Items for Systematic review and Meta-Analysis (PRISMA) guidelines,[12] and we registered the protocol on the International Prospective Register of Systematic Reviews (PROSPERO registration number CRD42021233862). Ethical approval was not required for this systematic review, since all data came from published articles.

### Effectiveness review

The effectiveness review was designed according to the PICO format as follows, through consultation with the WHO staff and expert group as part of the WHO guideline development process,[11] focusing on the aspect of self-monitoring versus clinic monitoring:

#### Population

Pregnant women and other pregnant people diagnosed with GD

#### Intervention

SMBG (either by the pregnant person or by another layperson, such as a family member) – Note: Although many products, devices, and mobile applications can be used to monitor blood glucose levels, we defined SMBG as home-based use of finger-prick devices, continuous glucose monitoring (including real-time), flash glucose monitoring, or urine dipstick for glucose testing.

#### Comparison

Clinic blood glucose monitoring by health workers during ANC contacts only

### Outcomes

Maternal:

1. Preterm labor
2. Caesarean section (including emergency C-section)
3. Long-term progression to type 2 diabetes or other metabolic disorders
4. Placenta previa
5. Hypertensive disorders of pregnancy (pre-eclampsia) or eclampsia
6. Self-efficacy, self-determination, autonomy, and empowerment
7. Device-related issues (e.g. test failure, problems with manufacturing, packaging, labelling, or instructions for use)
8. Follow-up care with appropriate management
9. Mental health and well-being (e.g. anxiety, stress, self-harm)
10. Social harms (including discrimination, intimate partner violence, stigma), and whether these harms were corrected/had redress available

Fetal/newborn:

1. Birth weight/size for gestational age (including macrosomia)
2. Respiratory distress syndrome
3. Stillbirth or perinatal death
4. Shoulder dystocia

### Inclusion criteria

To be included in the review, an article must have met the following criteria:

1. Study design that compared SMBG to clinic monitoring of blood glucose levels by health workers during ANC visits. This includes both randomized controlled trials, non-randomized controlled trials and comparative observational studies (including prospective controlled cohort studies, cross-sectional studies, controlled before-after studies and interrupted time series) that compare individuals who received the intervention to those who did not.
2. Measured one or more of the outcomes listed above
3. Published in a peer-reviewed journal

No restrictions were placed based on location of the intervention. No language restrictions were used on the search. Articles in English, French, Spanish, and Chinese were coded directly; articles in other languages were translated.

### Search strategy

The following electronic databases were searched through the search date of November 11, 2020: PubMed, CINAHL, LILACS, and EMBASE using the following search string (designed for Pubmed and adapted for the other databases).

(“glucose tolerance test”[Mesh] OR “oral glucose tolerance test”[tiab] OR “OGTT”[tiab] OR “blood glucose”[Mesh] OR “blood glucose”[tiab] OR “blood sugar”[tiab] OR “diabetes”[tiab] OR “gestational diabetes”[mesh] OR “gestational diabetes mellitus”[tiab] OR “glycemic index”[Mesh] OR “continuous glucose monitoring”[tiab] OR “glucose monitoring technique”[tiab] OR “glycemic control”[tiab] OR “flash glucose monitoring”[tiab])

AND

(pregnancy [Mesh] OR pregnancy [tiab] OR pregnant [tiab] OR peri-natal [tiab] OR perinatal [tiab] OR antenatal [tiab] OR maternal [tiab])

AND

(“self care”[Mesh] OR “self-care”[tiab] OR “self-monitoring”[tiab] OR “self-management”[tiab] OR “self-monitor”[tiab] OR “self-manage”[tiab] OR “self-monitored”[tiab] OR “self-managed”[tiab] OR “self-evaluate”[tiab] OR “self-evaluating”[tiab] OR “self-evaluation”[tiab] OR “self-test”[tiab] OR “self-testing”[tiab] OR “home”[tiab] OR “pharmacy”[tiab])

Secondary reference searching was conducted on all studies included in the review and relevant reviews.[13-18] We also searched for ongoing randomized controlled trials (RCTs) through clinicaltrials.gov, the WHO International Clinical Trials Registry Platform, the Pan African Clinical Trials Registry, and the Australian New Zealand Clinical Trials Registry. In addition, we searched the Cochrane Database of Systematic Reviews for potentially relevant articles cited in their reviews. Finally, selected experts in the field were contacted to identify additional articles not identified through other search methods.

Titles, abstracts, citation information, and descriptor terms of citations identified through the search strategy were screened by a member of the senior study staff. Full text articles were obtained of all selected abstracts and two independent reviewers assessed all full-text articles for eligibility to determine final study selection. Differences were resolved through consensus.

### Data management and analysis

Data were extracted independently by two reviewers using standardized data extraction forms. Differences in data extraction were resolved through consensus and referral to a senior study team member from WHO when necessary.

The following information was gathered from each included study:

- Study identification: Author(s); type of citation; year of publication
- Study description: Study objectives; location; population characteristics; definition of/diagnostic criteria for GD used in the study; type of blood glucose monitoring; description of self-monitoring access; description of any additional intervention components (e.g. any education, training, support provided); study design; sample size; follow-up periods and loss to follow-up
- Outcomes: Analytic approach; outcome measures; comparison groups; effect sizes; confidence intervals; significance levels; conclusions; limitations

For RCTs, risk of bias was assessed using the Cochrane Collaboration’s tool for assessing risk of bias.[19] For studies that were not randomized trials but were comparative, study rigor was assessed using the Evidence Project 8-item checklist for intervention evaluations.[20] Data were analyzed according to coding categories and outcomes. Where there were multiple studies reporting the same outcome, meta-analysis was conducted using random-effects models to combine risk ratios (RRs) or mean differences (MDs) with the program Comprehensive Meta-Analysis (CMA).

For each PICO outcome category, data were summarized in a GRADE Evidence Profile table using GRADEPro, prioritizing RCT data over observational data where available.

All analyses were stratified by the following categories/subgroups, where possible:

- Home monitoring (self, layperson, community health worker) vs. clinic monitoring outside of ANC (ambulatory, hospitalized, or additional to standard antenatal clinic visits)
- Type of glucose monitor
- Prior risk of (gestational) diabetes
- Vulnerabilities (i.e. obesity, age, poverty, disability, rural/urban, literacy/education level)
- High-income versus low or middle-income countries

### Complementary reviews

We conducted complementary reviews to examine the values and preferences of end-users and health workers and costs related to SMBG. We used the same search strategy as the effectiveness review to identify studies to be included in these reviews. These studies could have been qualitative or quantitative in nature, but had to present primary data collection; think pieces and review articles were not included. We summarized this literature qualitatively and organized findings by study design and methodology, location, and population.

#### Values and preferences review

We focused on studies examining the values and preferences of pregnant women and other pregnant people who were self-monitoring blood glucose levels or who were potential candidates for such self-monitoring, but we also included studies examining the values and preferences of health workers. We considered issues related to age of availability, informed decision-making, coercion, and seeking redress in this section; this included the effects of stock-outs or availability of glucose monitors.

#### Cost review

We included studies in this review if they presented primary data comparing costing, cost-effectiveness, cost-utility, or cost-benefit of the intervention and comparison listed in the PICO question above, or if they presented cost-effectiveness of the intervention as it related to the PICO outcomes listed above. This included both cost to the health system and cost to the end-user. Cost literature were classified into four categories: health sector costs, other sector costs, end-user/family costs, and productivity impacts.

## 2.3 Results

Our database search yielded 2787 records, and another 10 were identified through hand-searching and secondary searching (Figure 1). Of the 1871 unique records, we retained 78 for full-text review. Ultimately, we included 6 studies in the effectiveness review, 5 in the values and preferences review, and 1 in the cost review.

**Figure 1.**
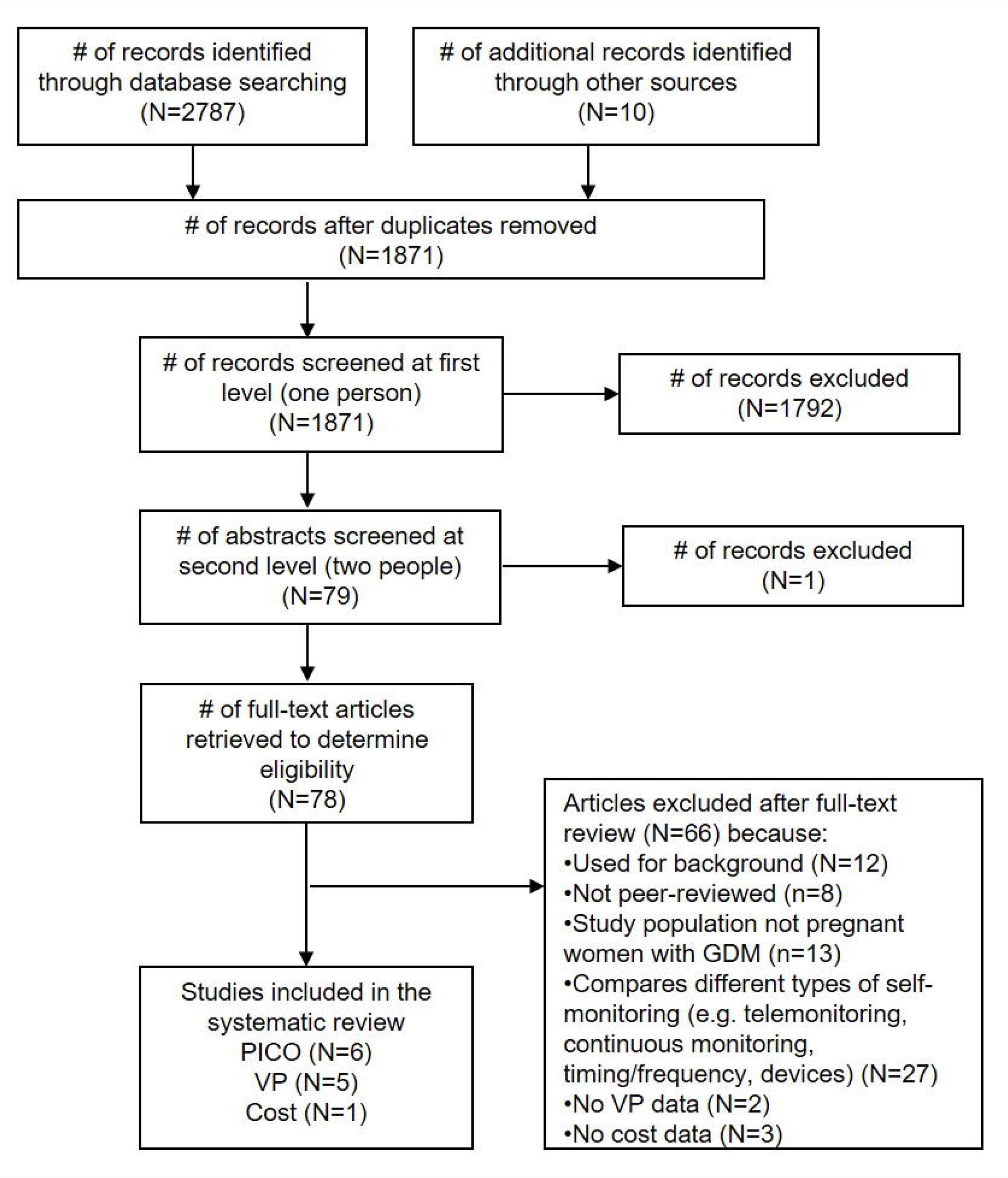
PRISMA flowchart for the search and screening process.

### Effectiveness review

For the effectiveness review, we identified 6 studies meeting the inclusion criteria: 3 RCTs and 3 observational studies.[21-26] The two larger RCTs (approximately 500 individuals per arm) compared SMBG as part of a package of interventions for GD treatment to routine care during antenatal contacts on clinical and healthcare utilization outcomes; one small RCT compared SMBG with periodic monitoring during prenatal visits on pregnancy and psychosocial outcomes (Table 1). While they did not specify the specific approach to glucose surveillance in the clinic setting, and while the results could not be disaggregated by intervention component, we opted to include these studies in the analysis as the closest available evidence for our PICO question. Both intervention and control groups ultimately received blood glucose monitoring and appropriate follow-up/treatment for GD; the difference was in self-versus clinic-monitoring. The 3 observational studies presented the same outcomes as the RCTs; therefore, to assess the highest-certainty evidence for each PICO outcome category, we included RCT data in the GRADE Evidence Profile (Table 2). Findings summarized in Table 2 represent pooled results from meta-analysis where multiple studies measured the same outcome, and the effect size of single studies where no other studies measured a specific outcome in a similar way. Meta-analysis results are presented in Figure 2. Given the small number of studies presenting outcome data, no further stratifications from our *a priori* list were possible.

**Table 1.** Description of included studies in the effectiveness review (RCTs)

| Study | Location | Population | Sampling | Intervention | Comparator |
| --- | --- | --- | --- | --- | --- |
| RCTs |  |  |  |  |  |
| **Crowther 2005 [22]** | Australia: 14 centers<br>UK: 4 centers | Women (24-34 weeks gestation) with GD, primiparous, singleton or twin pregnancy<br>Age (mean±sd): 30.9±5.4 (self); 30.1±5.5 (provider)<br>N=490 (self); 510 (provider) | Non-probability facility-based | Glucose self-monitoring (4 times a day) at home, as well as insulin therapy (if required), dietary advice from a dietitian, and ongoing care by attending obstetrical team with physician's support | Routine care at prenatal visits |
| **Landon 2009 [26]** | USA: 16 centers | Women (24-31 weeks gestation) with mild GD<br>Age (mean±sd): 29.2±5.7 (self); 28.9±5.6 (provider)<br>N=485 (self); 473 (provider) | Non-probability facility-based | Glucose self-monitoring (daily) at home using a portable memory-based reflectance meter, as well as formal nutritional counseling and diet therapy along with insulin (if required) | Routine care at prenatal visits |
| **Homko 2002 [25]** | USA: Philadelphia, PA | Women (<=33 weeks gestation) with GD<br>Age (mean±sd): 30.3±5.4 (self); 29.0±6.4 (provider)<br>N=31 (self); 27 (provider) | Non-probability facility-based | Glucose self-monitoring (4 times a day) at home using a reflectance meter with memory (One Touch Profile) | Routine care at prenatal visits |

**Table 2.**
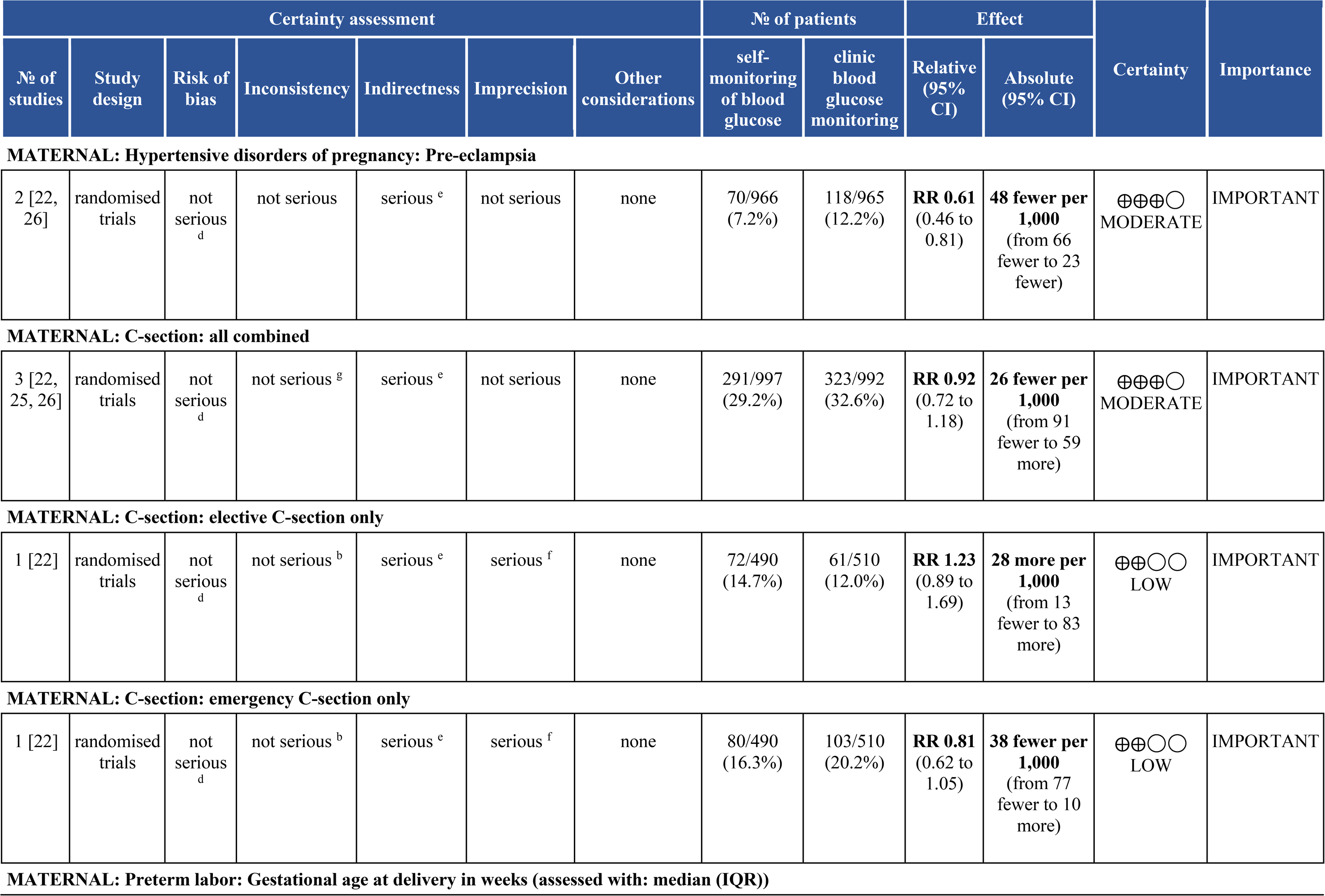
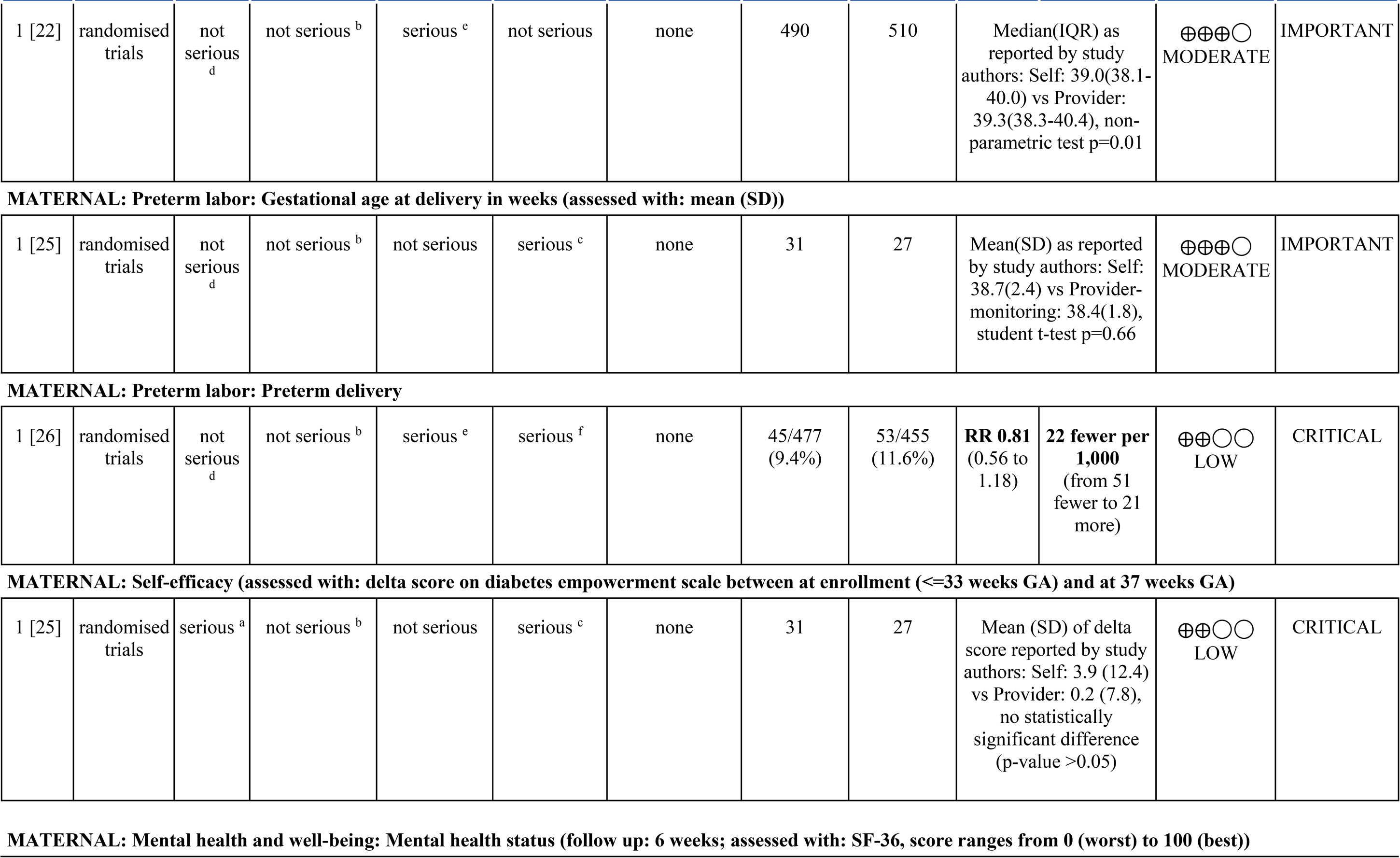
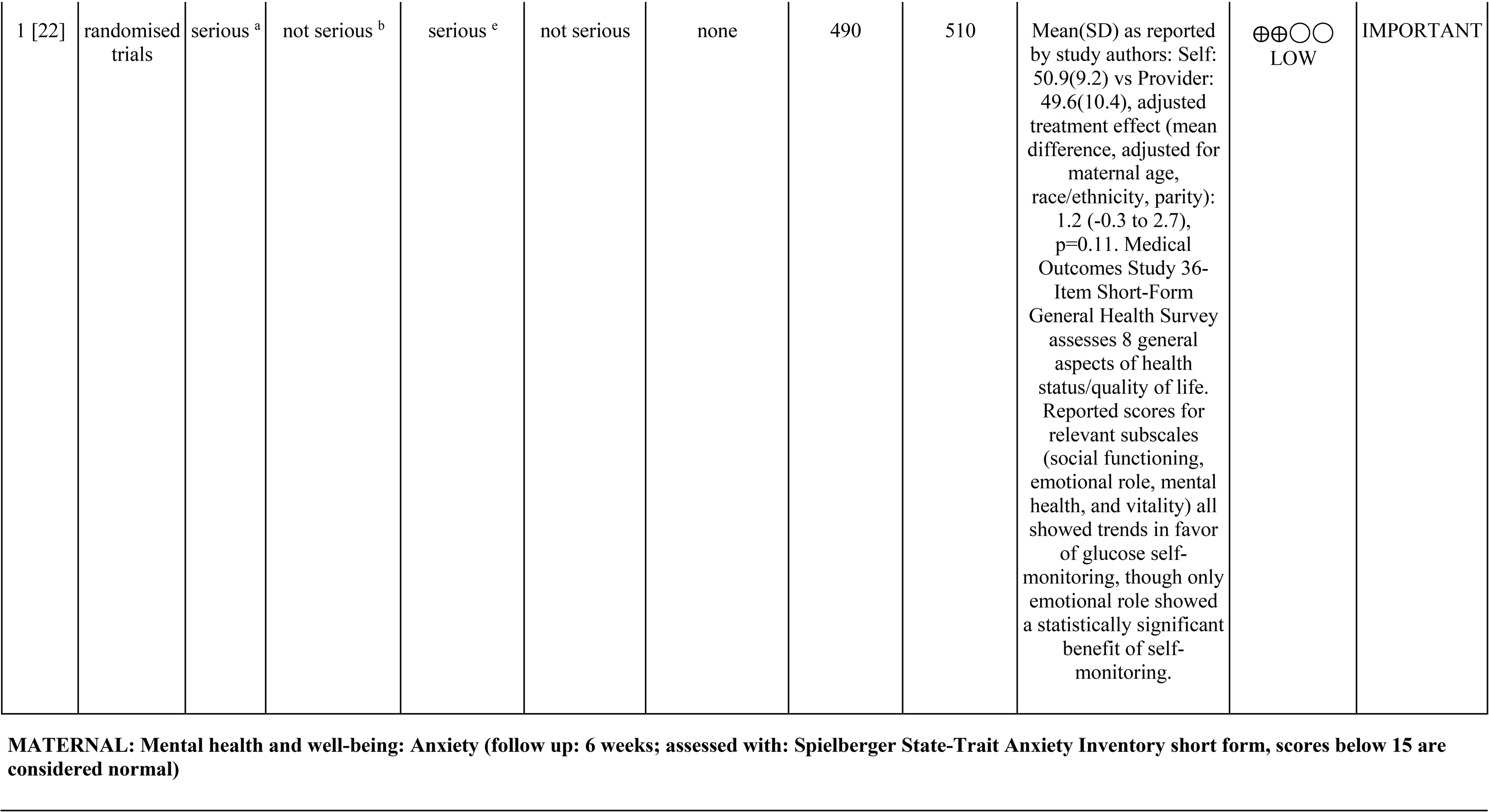
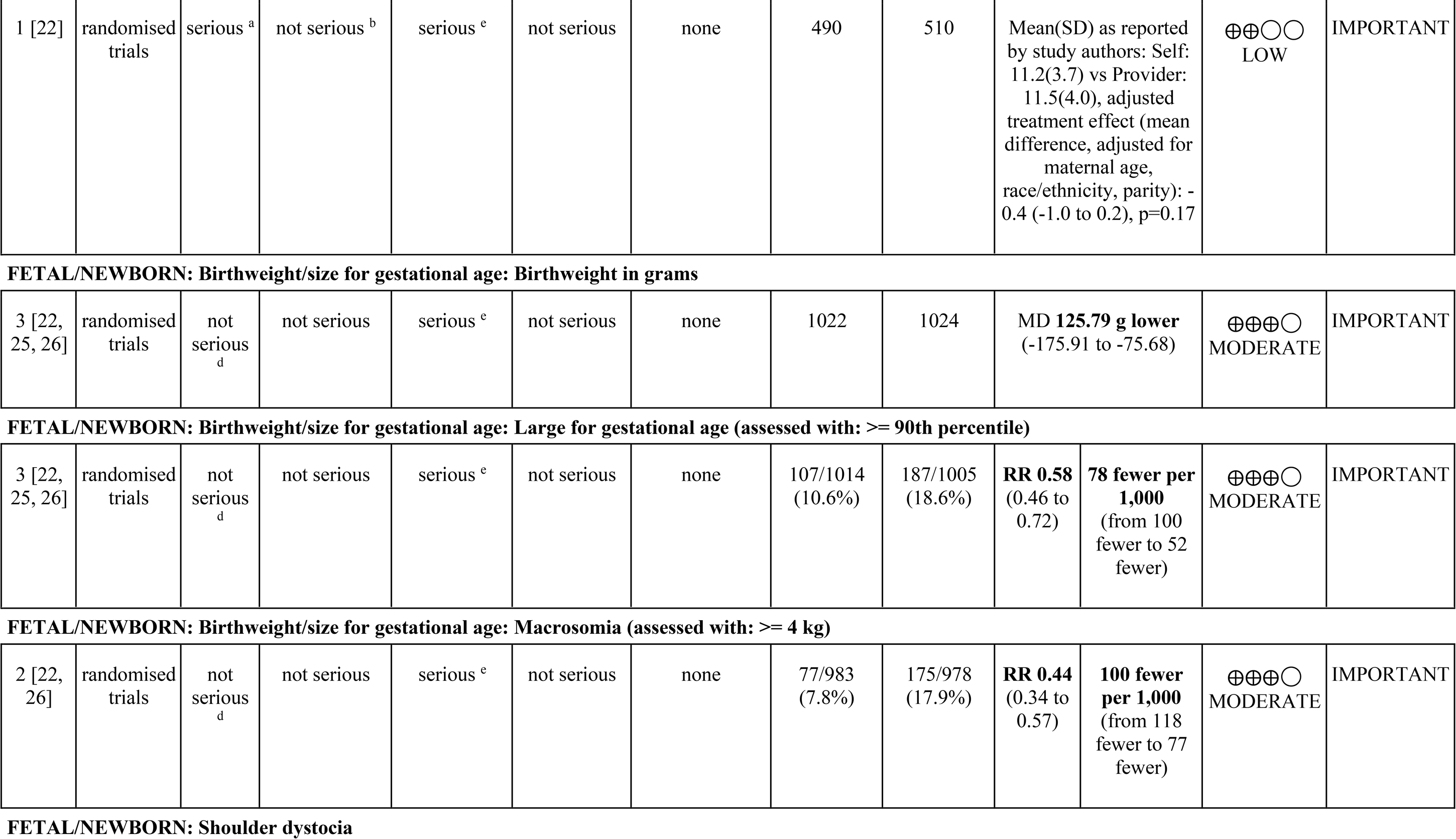
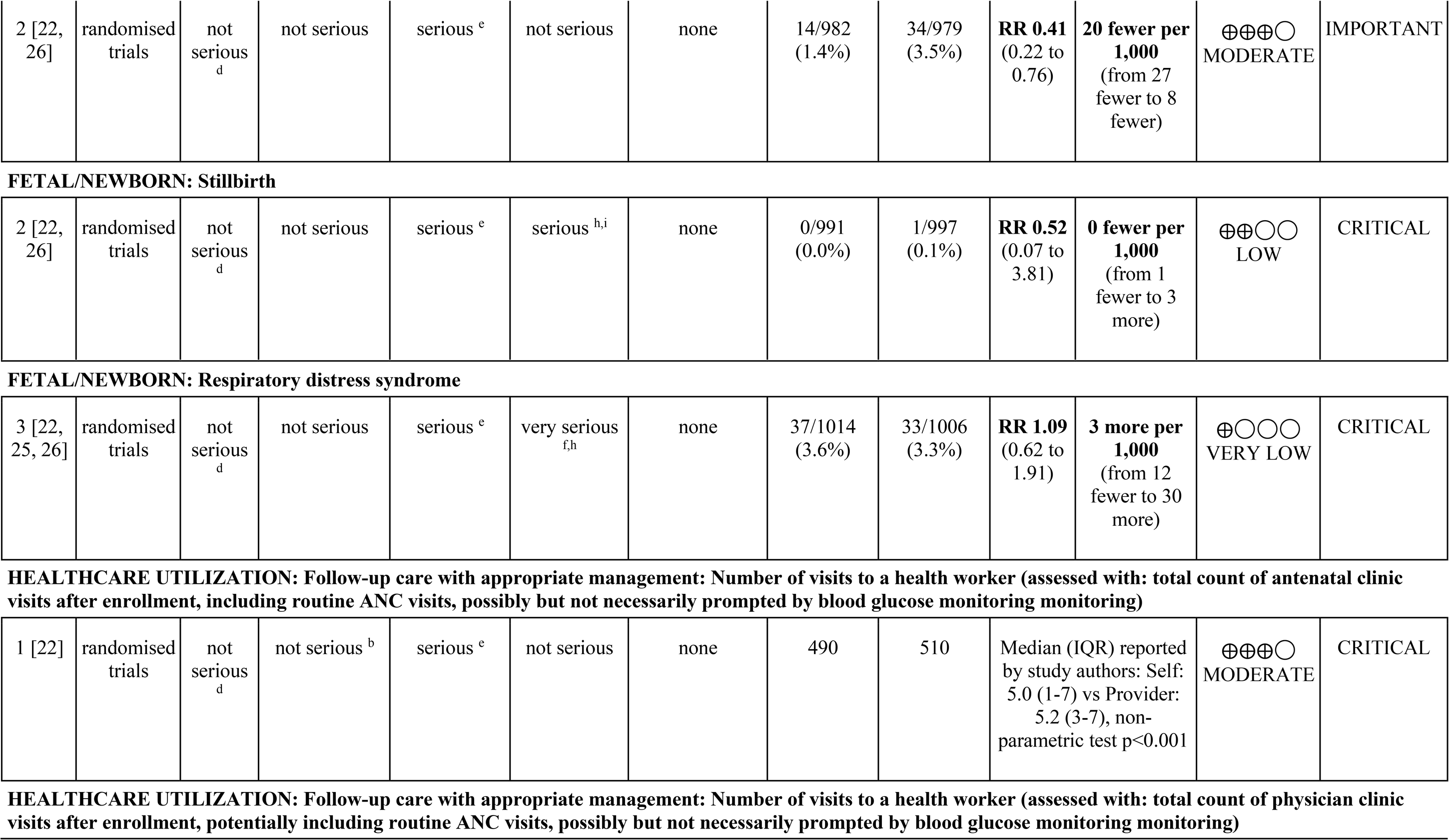
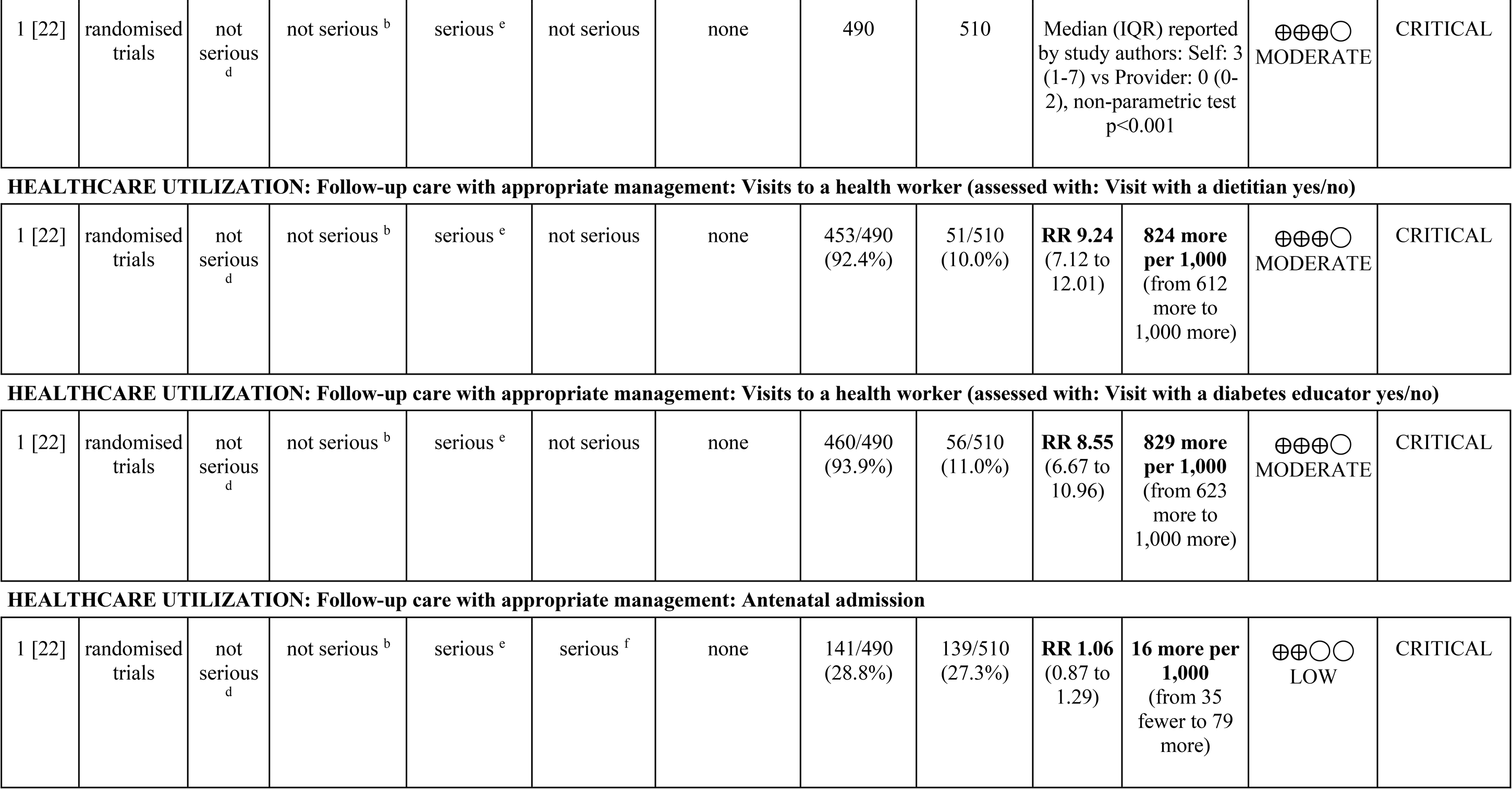
GRADE Evidence Profile.

**Figure 2.**
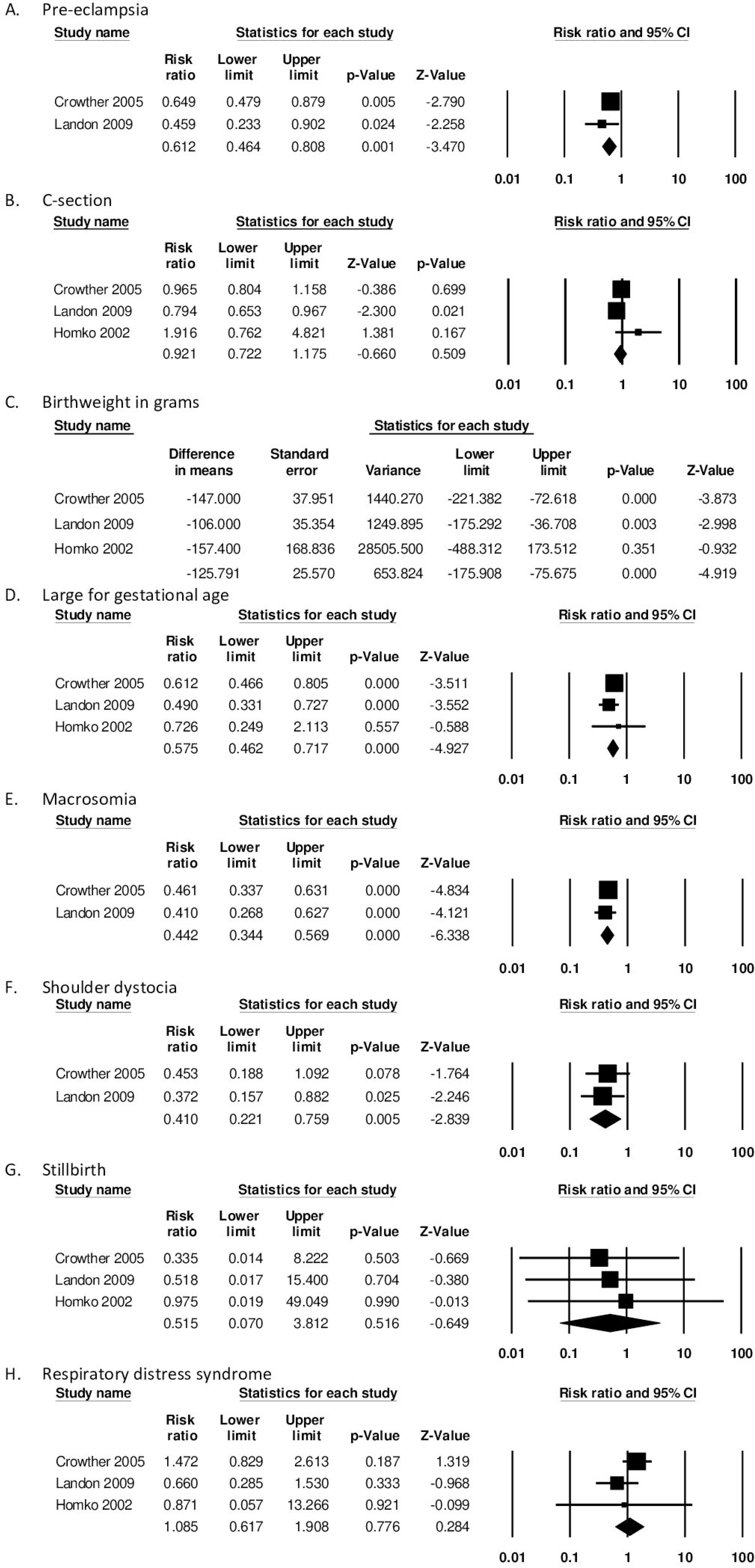
Forest plots and summary statistics from meta-analysis.

#### Maternal Outcomes

Moderate-certainty evidence from two RCTs demonstrated that SMBG as part of a package of interventions for GD treatment led to lower rates of preeclampsia (RR 0.61, 95% CI 0.46-0.81, Figure 2A).[22, 26] There was no difference on cesarean delivery rates (Figure 2B), with a pooled rate of 29.2% in the group that was treated, as compared to 32.6% in the untreated controls (RR 0.92, 95% CI 0.72-1.18), based on moderate-to low-certainty evidence from three RCTs.[22, 25, 26] One trial which disaggregated C-section outcomes by elective C-section and emergency C-section also found no difference between groups.[22] This package of interventions was not associated with gestational age at delivery[25, 26] or risk for preterm delivery[22] (RR 0. 81, 95% CI 0.56-1.18); this evidence was graded as low- to moderate-certainty.

In a small RCT in the United States, Homko and colleagues found no impact of SMBG as part of a package of interventions on self-efficacy based on self-empowerment score at 37 vs 33 weeks;[25] this evidence was graded as low certainty because of lack of blinding and the very small sample size. One RCT conducted in Australia and the United Kingdom showed SMBG as part of a package of interventions had no impact on validated questionnaire measures of mental health or anxiety.[22]

#### Fetal/neonatal Outcomes

Moderate-certainty evidence from 3 RCTs demonstrates that SMBG as part of a package of interventions was associated with changes in fetal growth, including lower mean birthweight (-126 g, 95% CI -176 to -76 g, Figure 2C) as well as lower risk for large for gestational age birthweight (RR 0.58, 95% CI 0.46 to 0.72, Figure 2D) and macrosomia (RR 0.44, 95% CI 0.34 to 0.57, Figure 2E) when compared to routine care during ANC contacts.[22, 25, 26] Two of these RCTs also demonstrated SMBG as part of a package of interventions was associated with lower rates of shoulder dystocia (RR 0.41, 95% CI 0.22 to 0.76, Figure 2F).[22, 26] There was no difference between groups for stillbirth rate[22, 26] (Figure 2G, low-certainty) or respiratory distress syndrome[22, 25, 26] (Figure 2H, very-low-certainty).

#### Healthcare Utilization

Crowther and colleagues quantified the impact of SMBG as part of a package of interventions for GD treatment on multiple measures of healthcare utilization.[22] In this study, both participants and health workers were blinded to the diagnosis of GD at randomization, and therefore treatment was, as expected, associated with more physician clinic visits and visits with dietitians and diabetes educators. Assignment to the treatment arm had no impact on antenatal hospital admissions.

No studies reported other quantitative comparative outcomes of interest for the effectiveness review, including long-term complications (such as both maternal and child type 2 diabetes, hypertension, or other metabolic disorders), device-related issues (e.g. test failure, problems with manufacturing, packaging, labeling, instructions), or social harms (e.g. discrimination, intimate partner violence, stigma).

### Values and Preferences Review

Five feasibility studies reported in 6 articles presented values and preferences data for specific blood glucose management systems (Table 3).[27-32] These studies (3 quantitative and 3 qualitative) all took place in high or upper-middle income countries: Canada, United Kingdom, Norway, Spain, and Thailand.

**Table 3.** Description of studies representing values and preferences data.

| Study | Location | Population and sample size | Data collection method | SMBG description |
| --- | --- | --- | --- | --- |
| Ardilouze et al., 2019 [27] | Canada | Pregnant women and other pregnant people ages 18+ years, with singleton pregnancy, 24-28-weeks gestation, 50 g GLT ≥7.2 mmol/L (GLT+), who were receiving pre- and perinatal care and did not have pre-pregnancy diabetes, disease or treatment with implications for glucose metabolism<br>N=103 | Questionnaires | Self-monitoring with a calibrated glucometer used after fasting and 2 hours postprandially, over 7 consecutive days without modifications to diet or exercise habits. Patients recorded their glucose levels on paper and transmitted them to the study nurse via e-mail or fax the day after the 7<sup>th</sup> day of measurements. They subsequently received questionnaires which were returned before diagnoses were announced to patients. |
| Garnweidner-Holme et al., 2018 and Skar et al., 2018 [28, 31]<br>Pregnant+ | Norway | Midwives (n=6) and nurses specializing in diabetes care (n=3) working at the diabetes outpatient clinic with a large population of women from diverse ethnic backgrounds; N=9 [28]<br>Pregnant women and other pregnant people of either ethnic Norwegian (n= 10) or immigrant backgrounds (i.e. born outside Norway and residing in Norway at the time of the study) (n= 7); N=17 [31] | Semi-structured interviews using the intervention group participants from a parent RCT (Pregnant+) | Self-monitoring by recording blood glucose levels accompanied by receiving verbal and written health and nutrition information, in line with clinic-based standard care protocol for GD.<br>The parent RCT added a use of a culture-sensitive mobile phone application to the standard care protocol. Patients could create their own profiles consisting of information related to but not limited to: their outpatient clinic, pre-pregnancy anthropometric measurements and physical activity level and preferred food culture. Patients could also record their physical activity time. Caregivers were requested to refrain from using the application to communicate as part of consultation. |
| Hirst et al., 2015 [29] | United Kingdom | Pregnant women and other pregnant people, with singleton pregnancy, whose GD was | Structured questionnaires | Self-monitoring using a Bluetooth-enabled glucometer (Polymap Glucose meter accessory with Lifescan UltraEasy meter) which automatically transmitted finger-prick blood |
|  |  | diagnosed prior to the 34<sup>th</sup> week of gestation, and having not received pharmacological therapy after 1 week of BG monitoring<br>N=49 |  | glucose level readings to a secure website via a smartphone application. Patients could label the readings with information regarding meals and medication use. The record was reviewed at least 3 times a week by caregiver team; patients and the team could communicate using the application. All equipment was loaned for free. |
| Rigla et al., 2018 [30] | Spain | Pregnant women and other pregnant people of greater than 19 years of age with GD diagnosed before the 34<sup>th</sup> week of gestation, having familiarity with smartphones and computer technology in general, and no history of pre-gestational diabetes, hemoglobin A1c >6.5%, active treatment with implications for changes in blood glucose levels, blindness, or severe mental disturbances<br>N=19 | Questionnaires | Self-monitoring using a smartphone with the clinical data collection and messaging system, including an accelerometer, Bluetooth-enabled glucometer (Accu-Chek Aviva Connect, Roche Diagnostics GmbH, Mannheim, Germany), Bluetooth-enabled blood pressure monitor (Bluetooth Blood Pressure Monitor 708-BT, Omron Healthcare Co, Ltd, Kyoto, Japan), and mechanism with which to report on the results of fasting daily ketonuria determination.<br>When it comes to the blood glucose level monitoring, patients were requested to download their readings (fasting and 1-hour postprandial) from the glucometer every 3 days. Patients were also requested to continue with their in-person visits to the clinic. |
| Youngwanichsetha et al., 2016 [32] | Thailand | Pregnant women and other pregnant people with GD diagnosed during 24- to 30-week gestation treated in the antenatal care units, diabetes clinics, or obstetric wards.<br>N=30 | Semi-structured interviews | Pregnant women and other pregnant people with GD were assumed to practice self-monitoring four times a day with a glucometer, as standard clinical practice in this hospital. The patients were to record the readings on paper or keep in the device memory. |

Overall, end-users found SMBG acceptable and even beneficial for a variety of reasons. Participants appreciated the technical convenience of using a smartphone for SMBG, which made recording and sharing blood glucose level readings easy,[29, 30] allowed for receiving feedback in real-time,[31] and kept important GD-related information handy as a resource.[31] Most believed that successful SMBG led to delivering healthy infants.[31, 32] However, this overall positive response appeared to be mostly from those who incorporated smartphone use in self-monitoring: one study which required participants use a glucometer and log book to record their blood glucose level values found only 6-7% of the surveyed end-users said that SMBG was convenient.[27]

Among end-users who self-monitored their blood glucose via smartphone, there was general consensus on the ability of SMBG to improve their confidence about health or self-care. Beyond finding SMBG useful and convenient, most stated they would recommend SMBG to other pregnant women and other pregnant people with GD.[30] End-users also found that SMBG increased their self-awareness and knowledge of their health status, amplifying their ability to effectively manage blood glucose levels during and after pregnancy.[31]

However, end-users also noted some concerns about SMBG. Some were frustrated with technical issues with the smartphone application: sometimes the application automatically transferred blood glucose values and registered wrong values.[31] Others expressed hesitation about the pain that comes with finger-pricking, though this dissipated over time and with experience.[32] When health workers lacked interest in the smartphone application, end-users were discouraged from continuing SMBG; most considered SMBG as a supplement to and not a replacement for usual ANC visits.[31]

One study in Norway reported values and preferences about SMBG from the health worker perspective.[28] Most participants agreed that SMBG through a smartphone application could help pregnant women and other pregnant people self-manage GM and found it useful for its convenience over paper-recording, especially given that modern technological progress would make app-based SMBG a more common practice over time. In addition, midwives and nurses reported liking the fact that the application could be resourceful for patients by providing helpful, credible health-related information to complement the SMBG records. However, some also expressed concerns that using the application alone may not allow patients to convey their emotion to their health care team, which could negatively affect the patient-provider relationship.

### Cost Review

No studies investigated the economic effects of SMBG in people with GD. One study reported economic effects of SMBG by patients with insulin-dependent diabetes during pregnancy (Orange County, California, USA).[33] Though this was a different study population than our population of interest, we used this study as indirect evidence for individuals with GD. Patients in the group using the reflectance colorimeter (SMBG) spent an average of 1.3 days in the hospital at a total average cost of US $593.00 as compared with the control group (conventional outpatient), who were hospitalized for an average of 3.8 days at an average of US $1,732.80. Only two of the nine patients in the meter group required admission as compared to five of the nine patients in the control group.

## 2.4 Discussion

This review attempted to answer the question of the value of SMBG for pregnant women and other pregnant people with GD. All three RCTs included in the effectiveness review compared SMBG as part of a package of interventions for GD treatment to routine care during ANC contacts. While they did not specify the specific approach to glucose surveillance in the clinic setting such that none of the comparison groups were explicitly aligned with the comparator in our PICO question but were approximations, and while the results could not be disaggregated by intervention component, the results highlight the value of SMBG as part of a larger program of treatment for GD. These studies showed that SMBG, in combination with other interventions for GD, was associated with maternal benefit, specifically lower risk of preeclampsia, as well as fetal benefits, including lower mean birthweight, fewer infants born large for gestational age, fewer infants with macrosomia, and lower rates of shoulder dystocia. In studies reporting end-users’ values and preferences, pregnant women and other pregnant people found SMBG acceptable and recognized benefits including convenience, ease of use, and increased confidence in managing their own health. Although we found no cost studies specifically on SMBG by individuals with GD, one study among pregnant women and other pregnant people with insulin-dependent diabetes found modest cost savings associated with SMBG.

Our findings must be interpreted in the context of limited available data. None of the effectiveness studies we identified included a control group with monitoring in the clinic setting, but rather had untreated “mild” GD receiving routine ANC. Inclusion of control participants with untreated GD likely exaggerates the impact of SMBG; however, we were unable to find any studies comparing SMBG to periodic monitoring in the ANC setting. While we hypothesize that isolated blood glucose monitoring in a clinic setting has limited utility, it is possible that periodic checks in the clinic setting have some benefit beyond no treatment at all given the likelihood of identifying the most overt hyperglycemia. However, participants with overt hyperglycemia on glucose screening tests were excluded from the RCTs included in this analysis.

In addition, though insulin therapy and dietary behavior modifications are both appropriate responses to the findings from blood glucose monitoring, because the included studies did not disaggregate data by the follow-up given after the monitoring (self vs clinic) step, we were not able to compare the effects of pharmacological intervention for GD in this review. Of the six included studies in the effectiveness review, five mentioned insulin therapy. Two of the three RCTs used in the effectiveness review listed in Table 1 compared SMBG (plus nutrition/diet counseling and insulin therapy if required) to routine prenatal care and did not disaggregate outcome effects by exposure to different components of the multi-component intervention,[22, 26] and the third RCT compared SMBG to clinic-monitoring in the context of diet-treated GD, though if a participant failed to meet metabolic targets they would start insulin.[25] Of the three observational studies, one study in the Czech Republic and another in Denmark compared SMBG to clinic-monitoring among women with insulin-treated GD,[21, 23] so both intervention and treatment groups received pharmacological intervention, and the third study in the USA compared SMBG to clinic-monitoring among diet-treated GD and explicitly excluded women who were initiated on insulin from the analysis.[24]

A strength of the review was the inclusion not only of effectiveness studies, but also of studies looking at costs and at values and preferences of patients and health workers. Costs to the patient, the health system, and society more broadly are an important consideration for any potential monitoring intervention. Potential drawbacks of SMBG as part of treatment of GD include increased healthcare utilization. One small study suggested potential cost savings; however, no studies examined out-of-pocket costs to individuals versus health system costs. Across multiple settings, values and preferences were generally positive towards SMBG, despite a few study participants noting inconveniences or frustrations with the technology/device. Studies generally pointed towards approval of expanded use of SMBG.

All of the studies included in the meta-analysis were conducted in high-income countries; only one values and preferences study was conducted outside of the United States and Europe (in Thailand). Health systems differ widely in their ability to provide care to individuals with GD, and data from a wider range of settings on effectiveness, values and preferences, and cost of this intervention would be valuable.

One concern that has been raised about SMBG is whether to conduct continuous glucose monitoring. While we included continuous glucose monitoring and intermittently-scanned (commonly known as Flash) glucose monitoring in our definition of SMBG, we excluded studies that compared different forms of SMBG, such as studies comparing continuous versus periodic SMBG. However, we note that a number of such studies have found SMBG positively associated with maternal and neonatal outcomes with continuous monitoring [34-36]; this approach has recently been recommended by some for GD [37]. Furthermore, in the context of the COVID-19 pandemic, possible delays in diagnosis and treatment could result in more advanced disease stages; delayed, incomplete or interrupted treatment and increases in behavioral risk factors, such as physical inactivity. Self-management actions are prioritized by WHO for maintaining essential NCD services during the pandemic.

Our review has several strengths. We used rigorous methods to search for, extract, grade and contextualize the evidence. We also included several outcomes beyond clinical pregnancy outcomes, including impact on maternal mental health and quality of life, as well as values and preferences and costs data. Together, these provide a more complete picture of the positive and negative aspects of this intervention. However, we did not include conference abstracts or grey literature, and the available peer-reviewed evidence was limited and came almost exclusively from high-income countries.

## Conclusions

SMBG during pregnancy among individuals with GD is feasible and acceptable, and when provided along with a package of interventions including insulin therapy, dietary counseling, and ongoing prenatal care with health workers, is generally associated with similar or improved maternal and neonatal health outcomes compared with standard care during ANC. However, more research is needed in resource-limited settings.

## Data Availability

Extracted data are available on request to the corresponding author.

## 3 Declarations

### 3.1 Ethics approval and consent to participate

Ethical approval and patients’ consent to participate was not required for this systematic review, since all data came from published articles.

### 3.2 Consent for publication

Not required, since all data came from published articles.

### 3.3 Availability of data and materials

Extracted data are available on request to the corresponding author.

## 4 Author Contributions

MN and OT conceptualized the study. CEK and PTY designed the protocol with feedback from OT, RG, BL, CZ and MN. PTY ran the database search and oversaw search, screening, full text review, and data abstraction processes with support from DR. CEK and PTY performed data analysis. PTY, CEK, and CZ drafted the manuscript. All authors reviewed the draft, provided critical review, and read and approved the final manuscript. The corresponding author, as guarantor, accepts full responsibility for the finished article has access to any data and controlled the decision to publish. The corresponding author attests that all listed authors meet the authorship criteria and that no others meeting the criteria have been omitted.

The named authors alone are responsible for the views expressed in this publication and do not necessarily represent the decisions or the policies of the World Health Organization (WHO) or the UNDP-UNFPA-UNICEF-WHO-World Bank Special Programme of Research, Development and Research Training in Human Reproduction (HRP).

The corresponding author, as guarantor, affirms that the manuscript is an honest, accurate, and transparent account of the study being reported; that no important aspects of the study have been omitted; and that any discrepancies from the study as planned (and, if relevant, registered) have been explained.

## 6 Acknowledgments

We thank Laura Ferguson, University of Southern California, and Maurice Bucagu, Department of Maternal, Child and Adolescent Health, World Health Organization, for their feedback on the review protocol and Bianca Hemmingson, Department of Non-Communicable Diseases, World Health Organization, for her feedback on the interpretation of the review findings. We also thank our research assistants from the Johns Hopkins Bloomberg School of Public Health (Jaime Marquis, Sarah Wagner, and Xuhao Yang) for their help in screening citations and abstracting data.

## Footnotes

1 While a majority of persons who are or can get pregnant are cisgender women, who were born and identify as female, transgender men and other gender diverse people may have the reproductive capacity to get pregnant. Therefore, the WHO guideline on self-care interventions which references the findings of this review uses language that is inclusive of all these experiences (“pregnant individuals”). In this manuscript, we use the term “pregnant women and other pregnant people” to include the preferred terminology of pregnant parents who use words other than women.

